# Cumulative seroprevalence among healthcare workers after the first wave of the COVID-19 pandemic in El Salvador, Central America

**DOI:** 10.1101/2022.02.06.22270565

**Authors:** Yu Nakagama, Maria-Virginia Rodriguez-Funes, Rhina Dominguez, Katherine-Sofia Candray-Medina, Naoto Uemura, Evariste Tshibangu-Kabamba, Yuko Nitahara, Natsuko Kaku, Akira Kaneko, Yasutoshi Kido

**Affiliations:** Department of Parasitology, Graduate School of Medicine, Osaka City University, Osaka, Japan; Research Center for Infectious Disease Sciences, Graduate School of Medicine, Osaka City University, Osaka, Japan; National Rosales Hospital, San Salvador, El Salvador; El Salvador National Institute of Health, San Salvador, El Salvador; Centro Nacional de Investigaciones Científicas de El Salvador (CICES), San Salvador, El Salvador; Department of Clinical Pharmacology and Therapeutics, School of Medicine, Oita University, Oita, Japan

**Keywords:** COVID-19, SARS-CoV-2, Healthcare workers, Seroprevalence

## Abstract

**Background:** The impact of novel coronavirus disease 2019 (COVID-19) on healthcare workers (HCWs) has been under-evaluated in Central America. We performed a seroepidemiological survey at a tertiary healthcare facility in El Salvador, where a large number of confirmed and far more suspected cases of severe acute respiratory syndrome coronavirus-2 (SARS-CoV-2) infected HCWs had been documented during the first wave of the pandemic.

**Methods:** During January–February 2021, a total 973 HCWs were tested for SARS-CoV-2 antibodies. Participants completed a questionnaire asking of their demographic data. Occupational risk was assessed by statistically comparing the seropositivity rates among different occupational categories.

**Results:** Overall seroprevalence in HCWs reached 52.6% (512 of 973). Of the seropositive individuals, 61.7% (316 of 512) had experienced a documented COVID-19 diagnosis, while the remaining 38.3% (196 of 512) were unrecognized seroconversions. Differences in seropositivity rates existed between occupational categories; nurses demonstrated the highest at 63.8% (222 of 348, risk ratio 1.44, p < 0.0001), followed by auxiliary HCWs assigned to patient-related work (55.9%, 52 of 93), and medical doctors (46.7%, 50 of 107). Several non-patient-related professions showed above-average seroprevalence, suggesting substantial SARS-CoV-2 contacts outside the workplace: 60.0% (6 of 10) and 68.0% (17 of 25) for nutritionists and pharmacists, respectively.

**Conclusions:** SARS-CoV-2 seroprevalence exceeded 50% among HCWs in El Salvador, with disparity among occupational categories with different workplace exposure risks. Importance of not only nosocomial infection prevention but also screening for transmissions having occurred outside the workplace were highlighted to efficiently control nosocomial spreads during a pandemic wave.

**Key points:** Healthcare workers in El Salvador were tested for SARS-CoV-2 antibodies. Seroprevalence reached 52.6%, with disparity among occupation; nurses ranked highest at 63.8% seropositivity. Alongside nosocomial transmissions, high seroprevalence associated with non-patient-related work suggested substantial SARS-CoV-2 contacts outside the workplace.

## Introduction

The novel coronavirus disease 2019 (COVID-19) pandemic has threatened the public health systems worldwide. Its impact, however, has been under-evaluated in most low- and middle-income countries, due to their lacking access to testing modalities. Central America is among the regions which utmost suffer scarcity of seroepidemiological data [1,2]. Under such circumstances, El Salvador reports a cumulative incidence rate at 1900 cases per 100,000 population by January 2022, ranking among the lowest in the Central Americas, only second to Nicaragua, where the government had officially set testing restrictions [3]. However, considering the limited testing capacity for active case detection, as well as further results from prior research reporting of excessive unrecognized seroconversions [4,5], substantial underestimation of the burden is anticipated.

Healthcare workers (HCWs) encounter the SARS-CoV-2 virus at the frontline and are representative of the highest risk individuals. Large healthcare-associated outbreaks not only endanger the assurance of quality care in the midst of a pandemic, but also may impact the kinetics of spread within the society [6]. We herein launched a seroepidemiological survey, extending from end-January to mid-February 2021, at a tertiary care referral hospital located in the capital city of El Salvador, where a large number of confirmed and far more suspected cases of SARS-CoV-2 affected HCWs had been documented during the first wave of the pandemic.

## Materials and Methods

### Study site, participants, data collection and sampling

A total 1144 HCWs from the National Clinical Laboratory and the National Rosales Hospital, El Salvador, were recruited to participate. The sites were the main referral facilities in the capital city of San Salvador for COVID-19 at the beginning of the pandemic from May to July 2020, having 116 beds across six wards constantly filled with patients of various severity. Strict measures were taken so that HCWs having been dedicated to work related to COVID-19 patients were isolated from the rest. Upon recruitment, the participants completed a questionnaire asking for their demographic data, anthropometric measurements, past medical history, occupational role, and clinical information related to any prior COVID-19 diagnosis given to the participant. Such clinical information included the mode of COVID-19 diagnosis (either laboratory-confirmed by polymerase chain reaction (PCR) based testing, or lacking a definitive laboratory test but clinically determined), self-reported signs/symptoms related with COVID-19, as well as their severity, and day of onset, if documented. Occupational roles were further divided into four categories: (i) medical doctors, (ii) nurses, (iii) other patient-related: those including the subcategories of respiratory care specialist, radiology personnel, cleaning staff, and other auxiliary HCWs who come into close contact with patients, such as transportation or security personnel, and (iv) non-patient-related: those who basically work aside from patients, including the subcategories of blood bank personnel, laboratory professional, administrative officer, nutritionist, and pharmacist. The presence/absence of a participants’ family member at his/her home with a COVID-19 diagnosis was queried to gain insight into the degrees of household contact/transmission. A total 973 subjects (973 of 1144, 85%), who consented to have their serum drawn and sent to Japan for evaluation, were eligible for the analysis (Figure). Sera were collected during a 3-weeks period (from end-January to mid-February 2021), aliquoted under sterile conditions, stored at −80°C, and shipped to Osaka City University, Osaka, Japan, where the serological analyses were performed.

**Figure.**
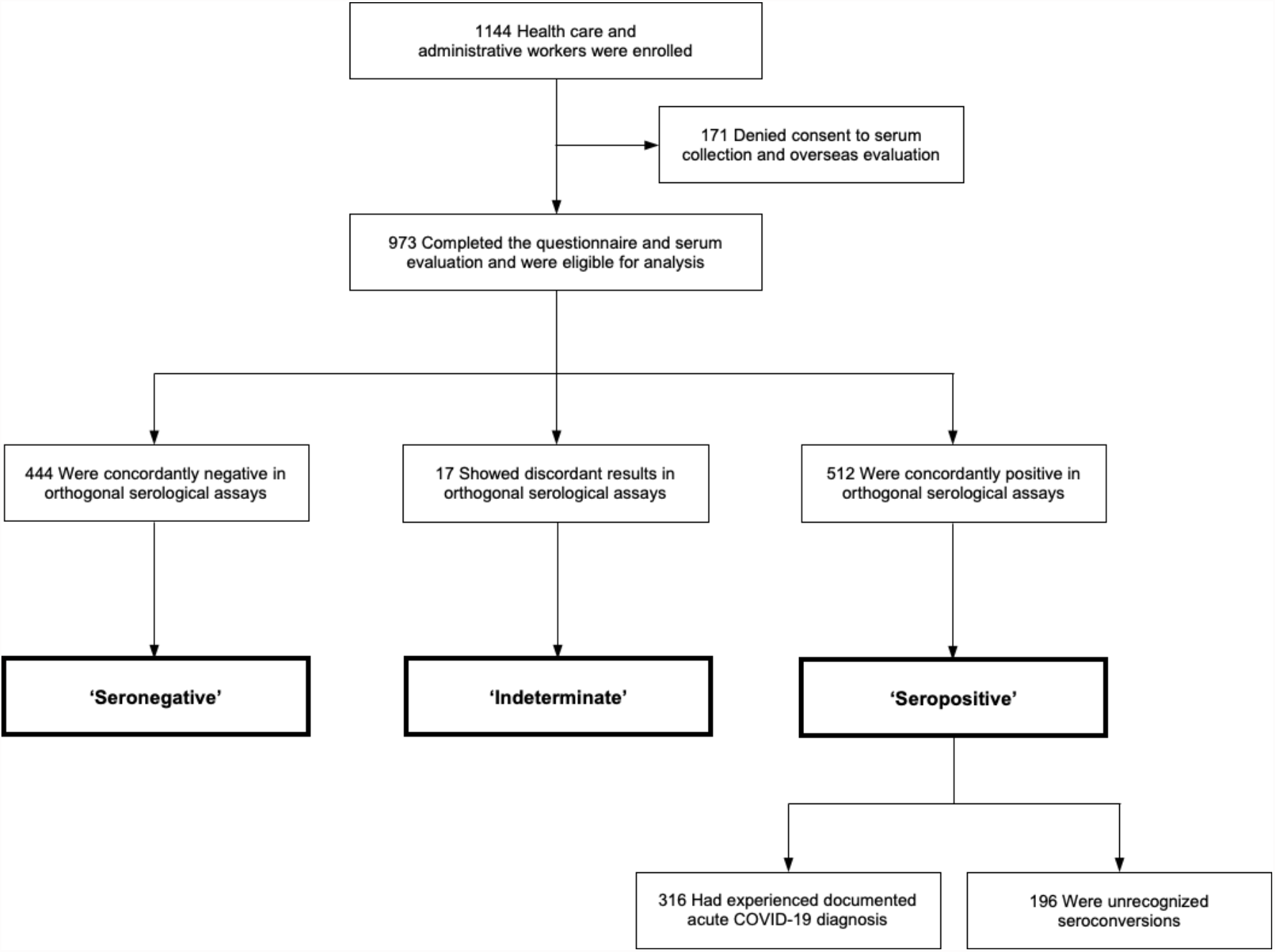
Enrollment, diagnostic algorithm, and results of testing.

### Ethical Statement

Analyses were conducted in accordance with the ethical standards noted in the 1964 Declaration of Helsinki and its later amendments. The research was approved by the Osaka City University Institutional Ethics Committee (#2020-003) and the National Research Ethics Committee of El Salvador (#CNEIS/2020/029). Consent for participation and publication was obtained from every participant.

### Serological testing and case definition

Two chemiluminescent immunoassays, the Roche Elecsys Anti-SARS-CoV-2 assay (Roche, Basel, Switzerland) and the Abbott SARS-CoV-2 IgG II Quant (Abbott, Chicago, Illinois, USA), were performed concomitantly, in accordance with the manufacturers’ instructions. The former targets the nucleocapsid protein (Roche) and the latter targets the spike receptor-binding domain (Abbott). A signal equal to or above the thresholds of 1.0 cutoff index (COI) and 50 AU/mL, respectively, were considered serologically positive. Serological status of an individual was defined based on the combination of results from the orthogonal testing. Participants were grouped as follows; (i) ‘seropositive’, if concordantly positive in both serological tests, (ii) ‘seronegative’, if concordantly negative in both serological tests, (iii) ‘indeterminate’, if discordant in the two immunoassay results. We have adopted similar orthogonal case detection approaches to successfully detect COVID-19 convalescent individuals with excellent sensitivity/specificity: 100% (95% CI, 68–100%) and 100% (95% CI, 94–100%), respectively [4,5].

### Statistical analysis

The results of serological testing were described as frequencies and percentages among the participants screened. Univariate analysis was performed to identify the variables associated with seropositivity. Comparisons of variables were made between ‘seronegative’ and ‘seropositive’ individuals. For the between-category comparison of seropositivity rate and the frequency of having a COVID-19 family member in the household, the ‘non-patient-related’ occupational category served as the reference group. Chi-square test was used for categorical comparisons of data. Mann-Whitney’s test was used to compare differences among continuous variables. Tests were two-sided, and a threshold of 0.05, or, for the multiple comparisons made between different occupational categories when assessing the differences in seropositivity rate and frequency of household contact, the Bonferroni-adjusted threshold of 0.017, was used to determine statistically significant p-values. Statistical analysis and artwork preparation were done using the GraphPad Prism software.

## Results

### Demographic description of the Salvadorian HCW cohort

A total 973 participants, aged 43 ± 11 years (mean ± SD), were enrolled (Figure, Table 1). Male participants constituted 28.3% (275 of 973) of the analyzed cohort. Participants were 11.0% (107 of 973) medical doctors, 35.8% (348 of 973) nurses, 9.6% (93 of 973) assigned to other patient-related work, and 43.7% (425 of 973) assigned to non-patient-related work. 25.3% (246 of 973) reported of having at least one high-risk chronic comorbidity, such as diabetes, hypertension, ischemic heart disease, cerebrovascular disease, cancer, and/or chronic obstructive pulmonary disease. Participants were 33.3% (324 of 973) obese, reporting a body mass index (BMI) of 30 or higher (Table 1).

**Table 1.**
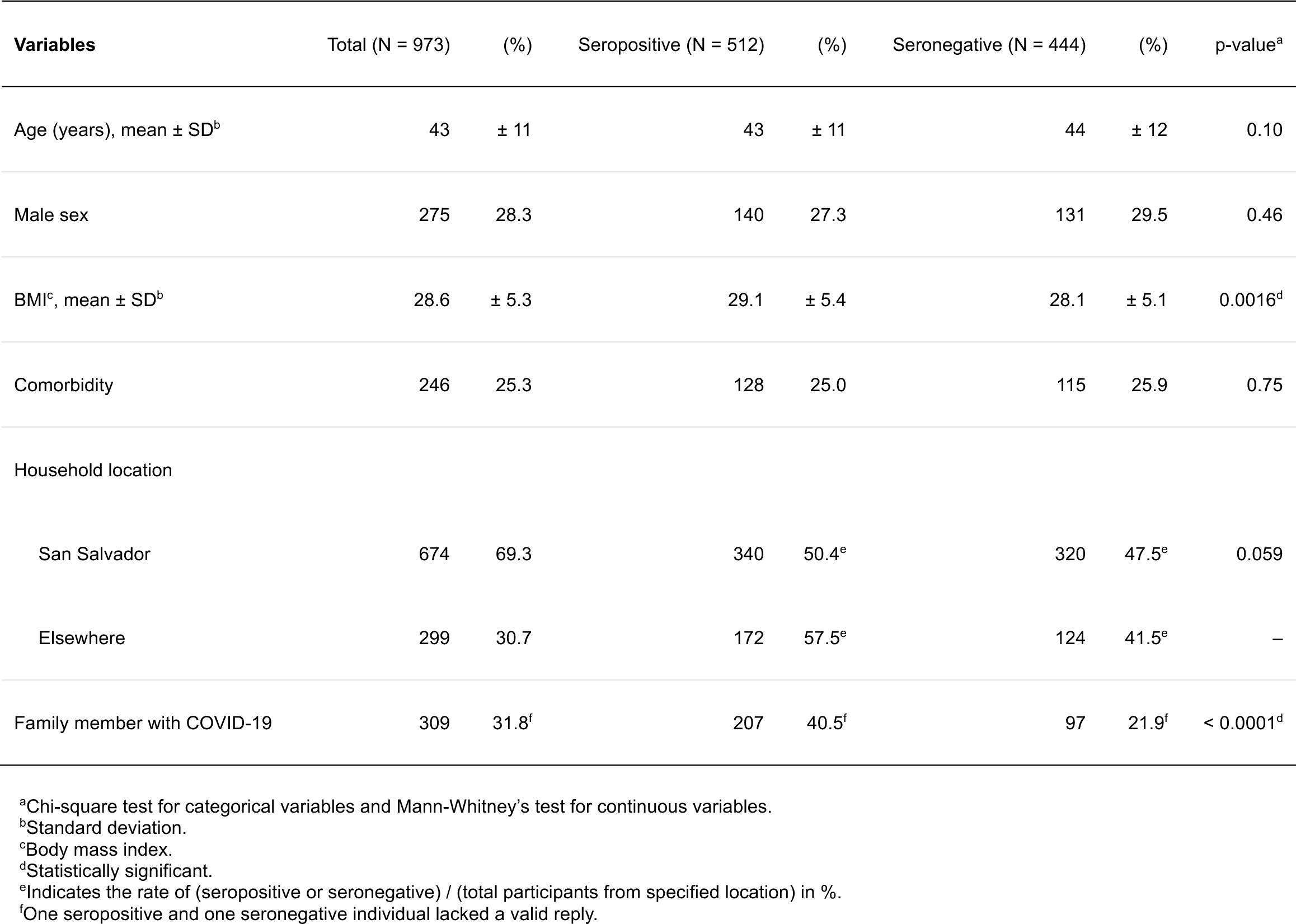
Factors related with SARS-CoV-2 seropositivity.

### Impact of SARS-CoV-2 among Salvadorian HCWs

The orthogonal testing algorithm revealed an overall seropositivity rate of 52.6% (512 of 973) among the HCWs of the National Rosales Hospital, San Salvador (Figure). Among the seropositive individuals, 61.7% (316 of 512) had experienced a documented COVID-19 diagnosis during the acute illness. The acutely diagnosed COVID-19 illnesses were 90.2% (285 of 316) symptomatic, and was laboratory-confirmed by PCR in 70.6% (223 of 316), while the remaining 29.4% (93 of 316) were clinically determined of COVID-19. Of the acute COVID-19 diagnoses, 7.0% (22 of 316) were treated with oxygen therapy, including one requiring mechanical ventilatory support. 5.9% (17 of 288) required hospitalization (A valid reply to the query on ‘hospital admission requirement’ was not available from 28 of 316 documented COVID-19 diagnoses). Finally, those never having been suspected of the diagnosis nor underwent SARS-CoV-2 PCR testing, but still revealed seropositive for anti-SARS-CoV-2 antibodies, aggregated to an excessive unrecognized seroconversion rate of 38.3% (196 of 512) among the total seropositive individuals.

45.6% (444 of 973) of the participants were concordantly negative for anti-SARS-CoV-2 serology. The remaining 1.7% (17 of 973) showed discordant results on the two immunoassays. Among the 17 individuals of indeterminate serological status, with discordant results on immunoassays, four (23.5%) had experienced symptomatic, laboratory-confirmed COVID-19 diagnoses, indicating a likely false-negative in one of the immunoassays.

### Factors related with seropositivity

In the further analysis, those with discordant serological testing results were excluded, and comparisons were made between ‘seropositive’ and ‘seronegative’ individuals to assess for factors related with seropositivity. Neither age in years (43 ± 11 vs 44 ± 12, p = 0.10) nor sex (male sex: 27.3% vs 29.5%, p = 0.46) were associated with serological status (seropositive vs seronegative, Table 1). Seropositive HCWs had slightly higher BMIs, of subtle clinical significance, compared with seronegative individuals (29.1 ± 5.4 vs 28.1 ± 5.1, p = 0.0016). The rate of carrying any comorbidity was comparable irrespective of serological status (25.0% vs 25.9%, p = 0.75). HCWs commuting from San Salvador, the department where the highest number of acute cases had been reported from [7], showed a trend towards lower seroprevalence compared with HCWs commuting from the other 13 departments of the country (50.4% vs 57.5%, p = 0.059), but the difference did not reach statistical significance.

Risk for seropositivity was next compared between different occupational categories (Table 2). Nurses had the highest seropositivity rate of 63.8% (222 of 348), followed by HCWs assigned to other patient-related work (55.9%, 52 of 93), and medical doctors (46.7%, 50 of 107). Of the HCWs assigned to other patient-related work, respiratory care specialists remained low in their seroprevalence (45.5%, 5 of 11), while cleaning staff showed a surprisingly high seropositivity rate (71.9%, 23 of 32). On the other hand, auxiliary HCWs assigned to non-patient-related work, as expected, exhibited the lowest seropositivity rate of all occupational categories (44.2%, 188 of 425). Interestingly, when subcategories of profession were further assessed, heterogeneity was observed in the seropositivity rate among HCWs assigned to non-patient-related work; nutritionists and pharmacists demonstrated above-average seropositivity rates of 60.0% (6 of 10) and 68.0% (17 of 25), respectively.

**Table 2.**
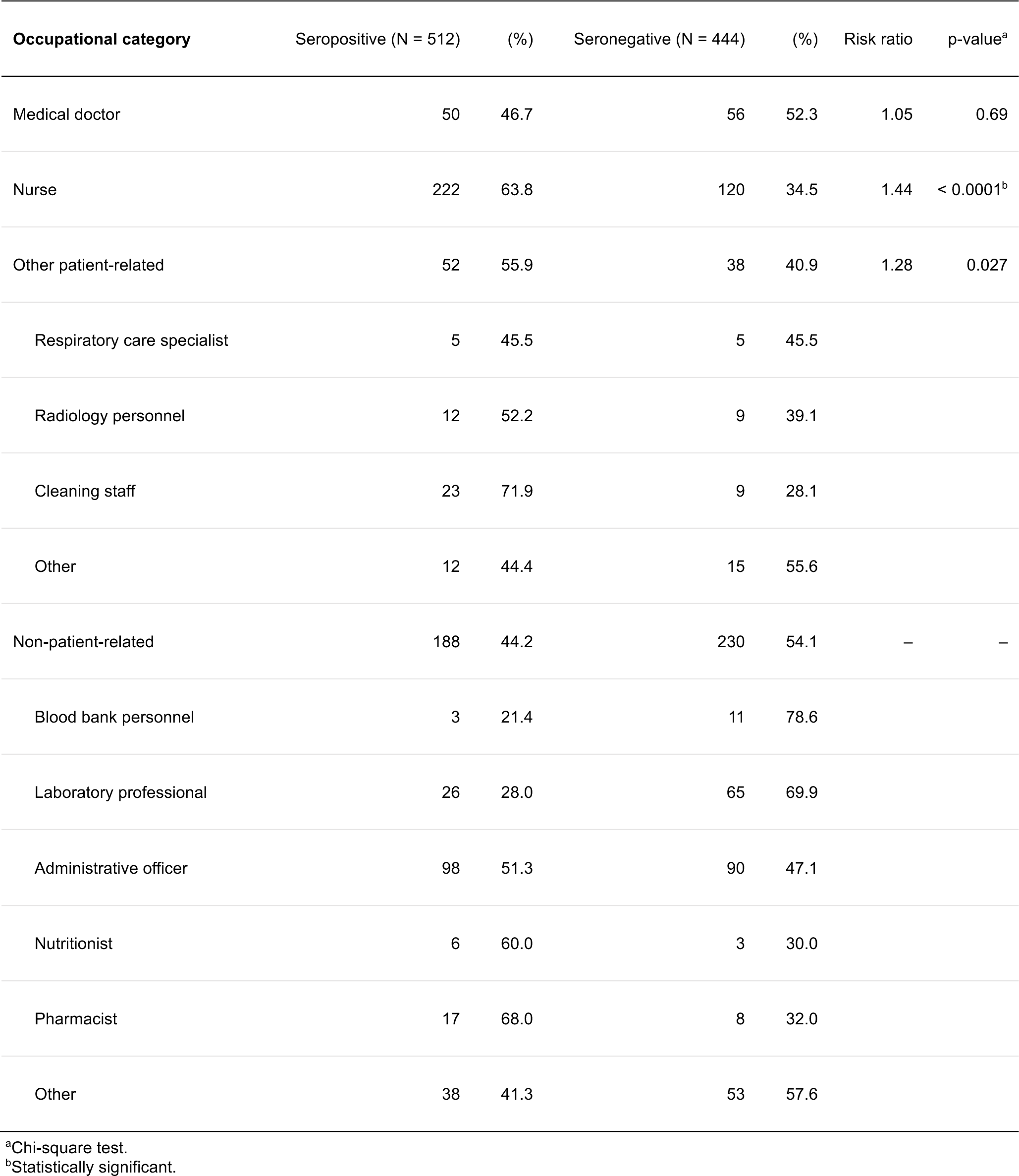
Difference in seropositivity rate between occupational categories.

Of the seropositive individuals, 40.5% (207 of 511) reported of at least one family member at their households with a COVID-19 diagnosis, the rate of which compared with seronegative individuals (21.9%, 97 of 443) was significantly higher (p < 0.0001, Table 1). The rate of reporting any family member with a COVID-19 diagnosis was almost uniform among occupational categories (Table 3). Such negligible variety in household contact rates was seemingly insufficient to explain the observed significant disparity in seropositivity rate among occupational categories.

**Table 3.**
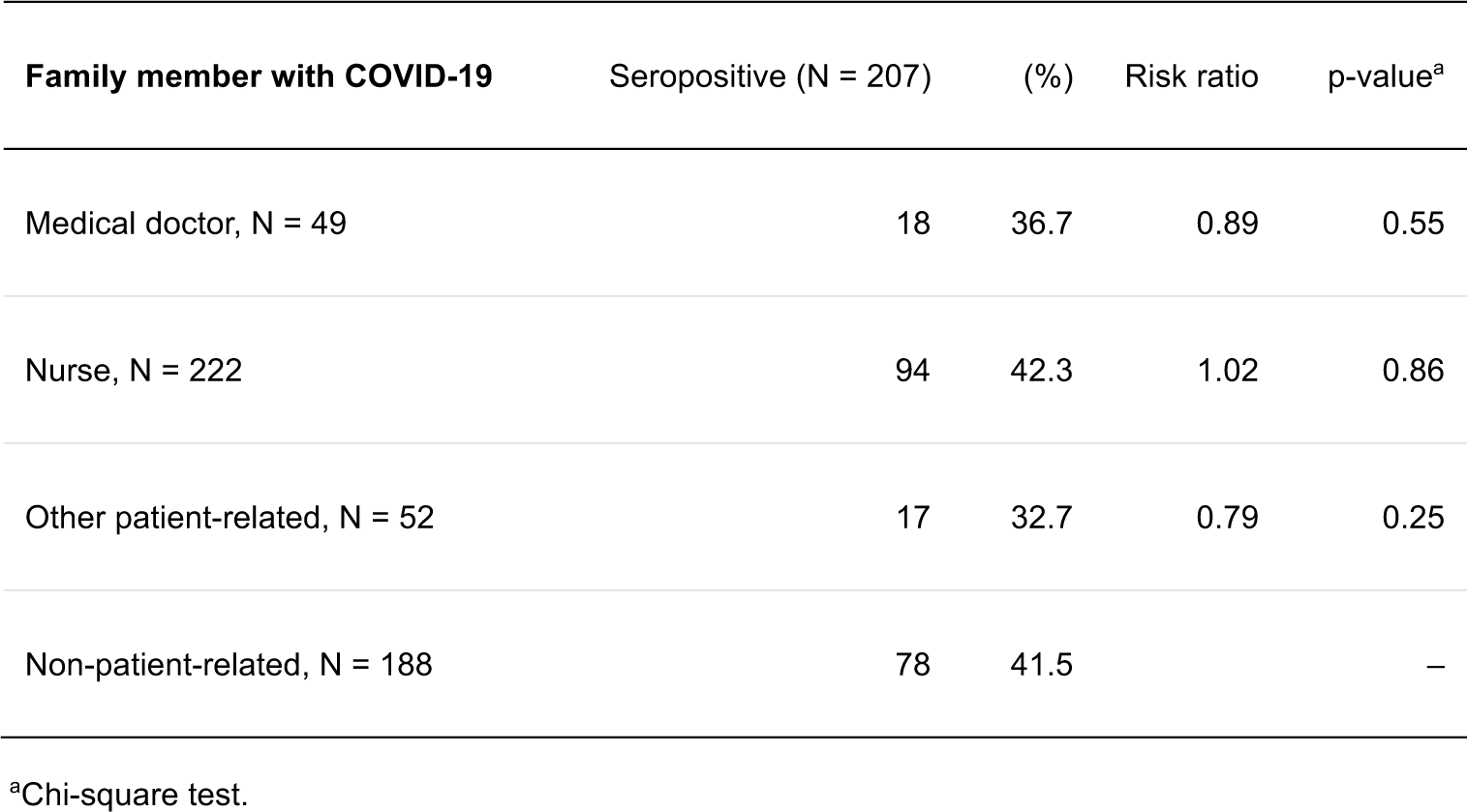
Frequency of household contact in seropositive individuals.

## Discussions

This is the first and only study to date, presenting data on SARS-CoV-2 seroprevalence in Salvadorian HCWs. In three independent meta-analysis studies, the overall seroprevalence of SARS-CoV-2 antibodies among HCWs was estimated to range globally from 8.0% to 8.7% [8–10]. One of the studies further described the regional differences in pooled serological prevalence of SARS-CoV-2 IgG antibodies; a higher seropositivity rate was observed in the USA (12.4%, 95% confidence interval (CI) = 7.8–17%) than in Europe (7.7%, 95% CI=6.3–9.2%) and East Asia (4.8%, 95% CI=2.9–6.7%) [8]. Our results, showing an overall seropositivity rate of 52.6% among a Salvadorian healthcare facility, listed at the highest level among previous publications on SARS-CoV-2 seroprevalence in HCWs. Furthermore, since serological tests are capable of exhaustively detecting past SARS-CoV-2 infections, including those never suspected of COVID-19 nor targeted for acute phase testing, an excessive unrecognized seroconversion rate of 38.3% (196 of 512) was unveiled among the total seropositive individuals. According to the epidemiological updates released by the Pan American Health Organization, the expected cumulative cases, as well as deaths, due to COVID-19 among HCWs in El Salvador had totaled to 6,609 cases and 71 deaths up until 8 February 2021, the date that approximates the period of conduct of our study [11]. Owing to the strengthening of the healthcare facilities’ preparedness, the rise of these numbers from El Salvador, as well as the entire Central Americas, have seemingly been curbed during the following months [12]. The significant number of unrecognized seroconversions revealed in the present study, however, highlights the possibility of these estimates being under-representative of the true burden.

The unique feature of the presented cohort was the discrepancy among occupational category in the risk of contracting the disease. Nurses, coming most frequently into direct and closest contact with COVID-19 patients, had significantly increased rates of seropositivity, compared with auxiliary non-patient-related HCWs. Conversely, risk differences were less evident between occupational categories in the studies targeting HCWs from Belgium, the United Kingdom or USA [13–15]. Taken together, the interpretation of occupational risk of workplace exposure imposed to frontline HCWs seems highly dependent on context such as national circumstances. The observed occupation-specific relative risks, that differ from report to report, are likely to reflect their local situations of practice, including access, as well as adherence to protective measures within the healthcare facility. The higher risk in the more near-patient HCWs shown in the presented cohort suggests that, despite strict protective measures set out for infection control, workplace transmission has persistently contributed to the spread of SARS-CoV-2 among Salvadorian HCWs. This may be attributable, at least in part, to the continuous shortage in material or technical support on the HCWs’ protective measures, which early on in the pandemic was a more universal situation [4,6]. Interestingly, when occupation was further divided into subcategories, there existed heterogeneous trends in seropositivity rates within patient-related, as well as non-patient-related professions. For example, respiratory care specialists in our HCW cohort, exclusively in charge of all tracheal intubation and extubation procedures of COVID-19 patients, carried less risk in contracting the disease compared with cleaning staff entering the COVID-19 ward. This apparently inverse relationship between aerosol exposure and seropositivity rate of the two subcategories, may have resulted from the differences in staffs’ preparedness for personal protection and infection control. Prior to the present study, Nicaragua and Panama were the only countries to have ever reported on the impact of SARS-CoV-2 among their HCWs [16,17]. Statistics on the mutual influence between the societal spread of SARS-CoV-2 and the healthcare systems’ preparedness in the Central Americas remain further awaited. Our observation gives initial insight into SARS-CoV-2 seroprevalence among HCWs in El Salvador, and shall serve as a reference guide for future seroepidemiological surveys to be performed in the country, as well as the neighboring Central Americas.

The impact of community transmission of SARS-CoV-2 in El Salvador is also yet to be elucidated, while the strictest lockdown measures undertaken, beginning from as early as 13 March 2020, promptly after the World Health Organization’s declaration of a global public health emergency, have surely minimized it. Within the non-patient-related HCWs, nutritionists and pharmacists were two subcategories having demonstrated above-average seropositivity rates. Harboring lowest risk in terms of workplace exposure, the rise in seroprevalence among several non-patient-related HCW subcategories may be attributable to such community transmission. Although not directly approached in the present study, the high frequency (21.9%) of seronegative individuals reporting of COVID-19 family members is highly indicative of substantial community transmission having taken place in El Salvador. The here observed bimodal pattern in seroprevalence among HCW of variable workplace exposure risks highlights the complexity and divergence of SARS-CoV-2 transmission routes affecting the healthcare setting. In addition to nosocomial infection prevention, screening HCWs for transmissions having occurred outside the working environment may serve as an efficient measure to control healthcare transmission during a pandemic wave.

Our study has several limitations. Firstly, upon estimating the impact of the pandemic on HCWs, we here adopted a rather conservative approach on case definition that shall minimize false-positive errors. Although this approach has performed well in previous seroprevalence surveys with excellent sensitivity/specificity, considering every individual with discordant immunoassay results as ‘seronegative’ may have led to false-negative interpretations and underestimation of seroprevalence in a high-prevalence cohort as the present one. The seropositivity rate was derived from a single institution survey and thus may not represent the general HCW population of the country. Secondly, our discussions regarding the site of SARS-CoV-2 transmission among HCWs, whether workplace or community, remains elusive due to the following constraints. The community’s seroprevalence data in El Salvador is still awaited in order to make direct comparisons with that of the HCWs. Transmission was also not directly monitored with molecular methods. Finally, the questionnaire was designed so as to limit the target of some selected queries; ‘presence/absence of COVID-19 symptoms’ were queried to those with documented infections. Collecting answers to this specific query exhaustively from every participant could have given us better estimates to the frequency of asymptomatic COVID-19 infections within the cohort.

In conclusion, we discovered that SARS-CoV-2 seropositivity rate exceeded 50% among HCWs from a referral hospital for COVID-19 in El Salvador. Significant differences were observed in the seropositivity rate among different occupational categories. Nurses were the most affected while high seropositivity rate was observed in several non-patient-related subcategories of profession. Alongside occupational exposures, protective measures are ought to target exposures outside the working environment with potential of subsequent introduction into the healthcare setting, in order to control healthcare transmission.

## Data Availability

All data produced in the present study are available upon reasonable request to the authors.

## Acknowledgments

The authors thank all healthcare workers who participated in the study as well as the co-investigators that facilitated the data collection and the material transfer needed for completion of the study.

